# Prevalence and Outcome of Cardio-Embolic Stroke Patients Admitted at Referral Neurology Hospital in Bangladesh

**DOI:** 10.1101/2022.02.16.22271069

**Authors:** S K Jakaria Been Sayeed, A K M Mahmudul Haque, Md. Moniruzzaman, Reaz Mahmud, Md. Abdullah Yusuf, Subir Chandra Das, Mohammad Bazlur Rashid, Sabrina Rahman, Abu Nayeem, A K M Humayon Kabir, M S Jahirul Haque Chowdhury, Md. Mujibur Rahman

## Abstract

**Background:** Stroke is the second leading cause of mortality worldwide; where the majority of stroke is ischemic. Among ischemic stroke, cardio-embolic has both higher severity and mortality.

**Objective:** To find out clinical outcomes and determine predictors of mortality related to cardio-embolic stroke.

**Methodology:** This prospective cohort study was conducted among patients of acute ischemic stroke of cardiac origin admitted at the National Institute of Neurosciences and Hospital, Bangladesh from 1^st^ October 2020 to 30 September, 2021. Patients were kept under follow-up to 90 days from discharge.

**Results:** A total of 689 ischemic stroke patients were screened, 156 had confirmed Cardio-embolic stroke. So, the frequency of cardio-embolic stroke was 22.64%. Male to female ratio was 1.3:1, mean age of 63 years. Hypertension 119 (76.3%), atrial fibrillation 107 (68.6%), and IHD 40 (25.6%) were most common comorbidities. Interestingly, we found only 23 (14.7%) patients with chronic rheumatic heart diseases. NIH Stroke scale score (median, IQR) during admission was 13 [7-19]. Overall mortality was 47 (29.9%), among them 30 (19.2%) died within 48 hours of hospital admission while 17 (10.9%) within 90 days of hospital discharge. Modified Rankin score at 90 days was 2 [min 0, max 5] those who survived. Cumulative incidence of recurrent stroke was 9 (7.1%) and incidence of anticoagulant induced hemorrhage were 5 (3.2%) among them. Risk factors associated with mortality (odds ratio, [95% CI], p value) were acute myocardial infarction (1.6 [1.14 – 2.52], 0.04), raised Troponin (1.89 [1.16-2.99], 0.01), reduced ejection fraction (3.38 [2.17-5.27], <0.001), hypotension (3.12 [2.07 – 4.68], < 0.001), chronic kidney disease (1.82 [1.06 - 3.10], 0.04), raised Creatinine (2.41 [1.52 - 3.84], 0.01), raised blood sugar (1.82 [1.14 - 2.89], 0.02), severe stroke (9.45 [3.57 – 25.03], <0.001), large infarct (53.67 [7.59 - 379.47], < 0.001), hemorrhagic transformation (4.43 [2.89 – 6.84], < 0.001) and aspiration pneumonia (1.9 [1.28-2.39], 0.01).

**Conclusion:** Overall frequency, severity, functional disability, and mortality in cardio-embolic stroke are higher. Acute myocardial infarction, severe stroke, presence of hyperglycemia, hypotension, renal impairment, low ejection fraction, large infarct, hemorrhagic transformations, and aspiration pneumonia are both clinically and statistically significantly associated with mortality in cardio-embolic stroke.

## Introduction

Each year twenty-six million people worldwide experience a stroke and it is the second-leading cause of mortality and a leading cause of long-term disability [1]. One-third of strokes represent intracerebral or subarachnoid hemorrhage, whereas two-thirds represent cerebral ischemia [1]. The reported prevalence of stroke in Bangladesh is 0.3 −1% [2, 3]. Approximately one in four ischemic strokes is of cardio-embolic origin [4]. Atherosclerosis of the cerebral circulation, occlusion of cerebral small vessels, and cardiac embolism are the major causes of ischemic stroke [5]. According to Stroke Data Bank and Registries divided the potential cardiac causes of the stroke into strong sources (prosthetic valves, atrial fibrillation, sick-sinus syndrome, ventricular aneurysm, akinetic segments, mural thrombi, cardiomyopathy and diffuse ventricular hypokinesia) and weak sources (myocardial infarct in earlier months, aortic and mitral stenosis, aortic, and mitral regurgitation, congestive heart failure, mitral valve prolapse, mitral annulus calcification, and hypokinetic ventricular segments) [6, 7]. Cryptogenic stroke accounts for 25% of all ischemic strokes, the majority of which is likely to be of embolic origin [8]. Cardio-embolic stroke has been increasing in number not only in the developed country but also in low to middle-income countries whereas the overall incidence of stroke is decreasing due to adequate treatment and prevention strategies against hypertension, dyslipidemia [9, 10, and 11]. In Bangladesh, chronic rheumatic heart disease is one of the common causes of cardio-embolic stroke, prevalence of rheumatic heart disease is 0.9 per 1000 [12]. The risk of recurrence in cardio-embolic stroke, is highest (around 10%) in the first weeks after the stroke that drops to 5% in the following 12 months, however, the risk of early embolic recurrence varies between 1 to 10% [13]. Cardio-embolic stroke is known to cause more severe stroke and higher mortality [14] than other stroke subtypes. However, in Bangladesh prevalence of cardio-embolic stroke is not adequately evaluated, there is a lack of sufficient information regarding clinical & laboratory characteristics. Moreover, information regarding neurological and cardiac outcomes is still not known. Nevertheless, effect & outcome of anticoagulant therapy especially in myocardial infarction, rheumatic heart disease, or atrial fibrillation with ischemic stroke in our country still inadequately described. For this reason, we have conducted this prospective study to describe hospital prevalence, clinical and anticoagulant treatment outcomes among patients who were admitted to the National Institute of Neurosciences & Hospital. It is obvious; the study result will help the clinicians as well as policy makers to identify the original statistics and plan of management of this group of patients.

## Materials and Methods

### Study Settings and Populations

This was a prospective study, conducted at the National Institute of Neurosciences, the largest neurology referral hospital in Bangladesh. All acute ischemic stroke patients due to cardio-embolism admitted for the first time in NINS were enrolled [Figure 1].

**Figure 1:**
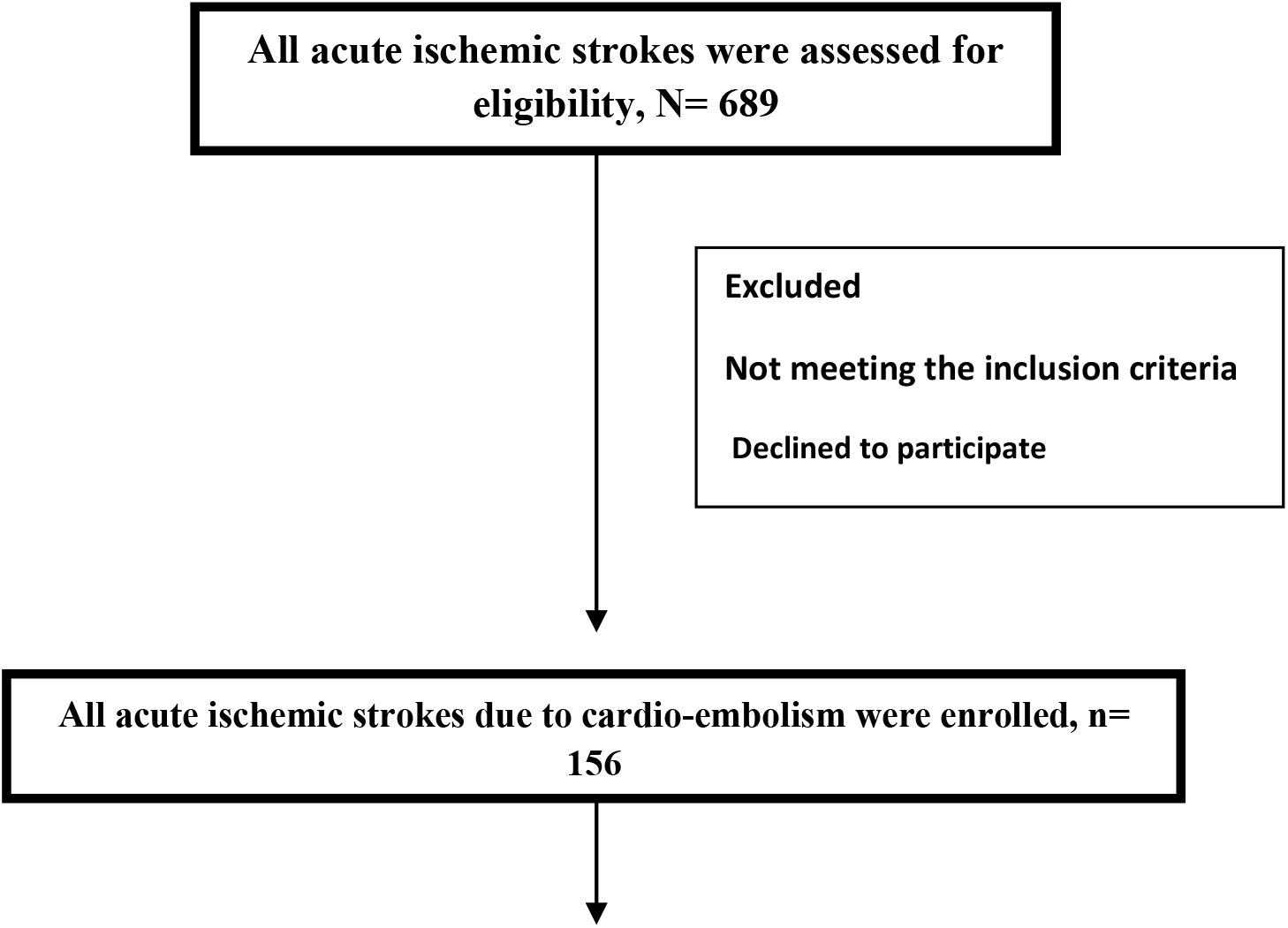
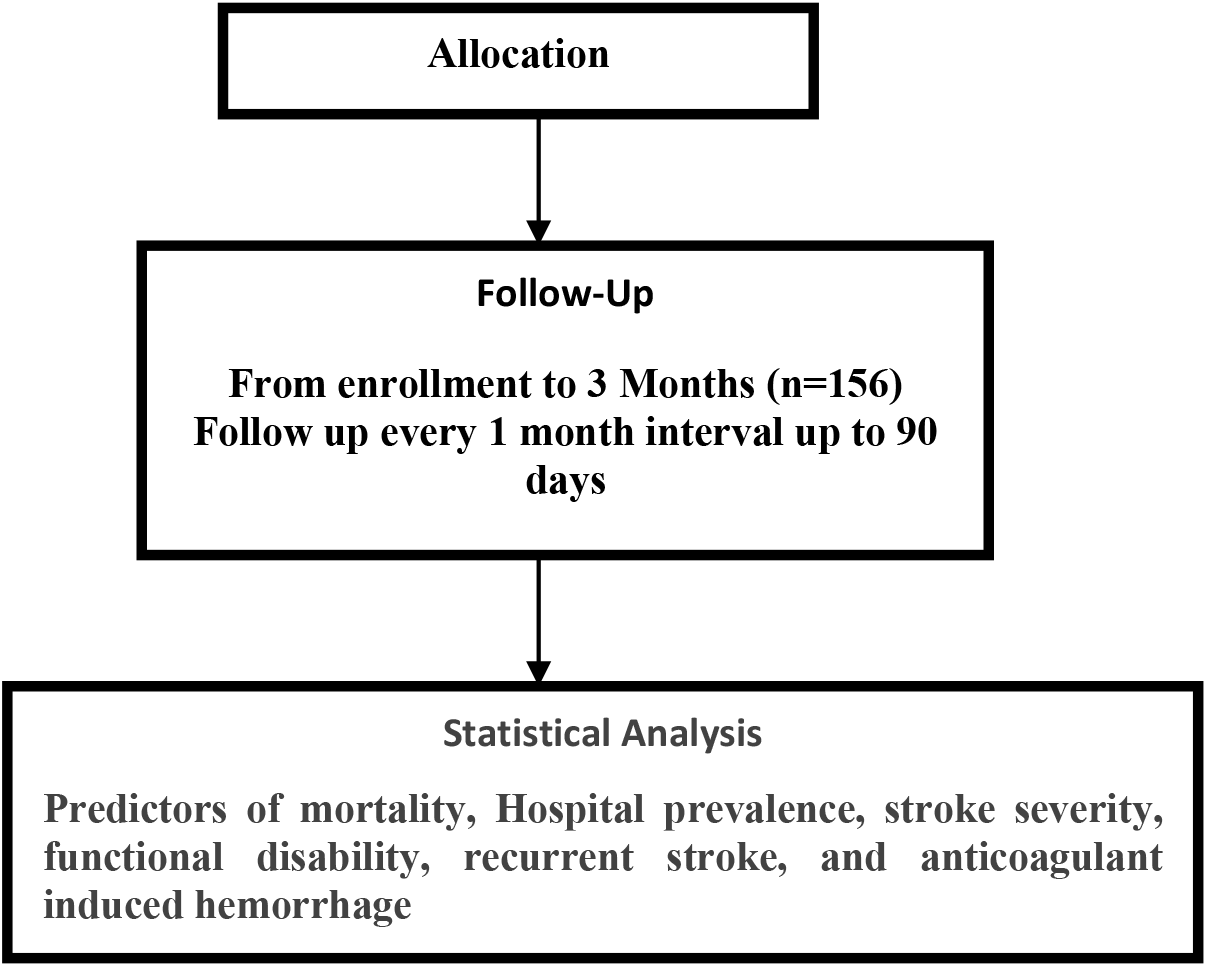
Schematic Flow Chart Of Research Work

### Inclusion criteria

Patients, more than 18 years of age with acute ischemic stroke diagnosed by CT or MRI within 1 week of index events due to cardiac causes diagnosed by electrocardiogram, echocardiography, (Arrhythmia, Acute MI, CRHD, Left ventricular aneurysm, prosthetic heart valve, atrial myxoma or any thrombus or vegetation in the cardiac chamber or valve surface) was included in the study.

### Exclusion criteria

Patients with intracranial hemorrhage or infection or tumor, transient ischemic attack, ischemic stroke due to neck vessels thrombus/ disease, severe organ dysfunction (renal and liver Injury), and other causes of cerebral infarction (Coagulopathy, Vasculitis, and tumor) were excluded.

### Outcomes Measure

Primary outcome was to determine predictors of mortality at 90 days after the index event. Secondary outcomes were assessing functional status by modified ranking scale score (mRS), recurrent stroke (ischemic or hemorrhagic) at 90 days after the index event. Moreover, we also determined the hospital frequency, measured stroke severity by using NIHSS (National Institute of Health Stroke Score) during admission and before discharge, and anticoagulant induced hemorrhage especially intracranial hemorrhage at 90 days.

### Investigations & Follow up

For, each patient’s base line investigations like CBC, Urine R/E, Creatinine, SGPT, RBS, HbA1C, Electrolyte with special investigations like (Coagulation profile, D-dimer, Troponin I, Fasting lipid profile, TSH, ANA, ANCA if needed) were done. 12 lead ECG, 24 hours halter (if applicable), Echocardiography (2D, M mode, color Doppler), CT/ MR angiogram of Brain and Neck vessels were done where necessary. Here, all investigations were done at NINS. We kept discharged patients under regular follow-up for up to 90 days from the index event. After discharge 1^st^ follow-up was given 1 month from index event, thereafter 2^nd^ follow-up 1 month from 1^st^ follow-up, again 3^rd^ follow-up was given 1 month from 2^nd^ follow-up. As the COVID situation is ongoing we had maintained follow-up using social media like what’s app, messenger, imo and viber (video conference) whatever available. Two trained doctors were allocated for data collection, follow-up schedule maintenance. Those who suffered a further stroke were admitted in the same institute for evaluation and management.

### Statistical Analysis

As cardio-embolic stroke prevalence in Bangladesh is 4.9% according to Bhowmik NB et al. [15]. Therefore, we assume 72 will be the size according to the prevalence equation. However, we enrolled 156 cardio-embolic stroke patients among 689 acute ischemic stroke patients as per inclusion criteria. The Sampling technique was purposive & consecutive. All data had been analyzed in SPSS 24 version. Continuous variable had been expressed with number, mean and standard deviation (SD) while value with skewed deviation expressed as IQR (Interquartile Range). Comparison between groups (Group A-alive, Group B-death) with mortality & functional outcome were analyzed by Pearson chi-square test for categorical variable. For continuous variable independent student’s t (normally distributed) or Mann Whitney U test for skewed data were applied. For assessing predictors of mortality, binary logistic regression analysis was done. Predictors were expressed with odds ratio with 95% confidence interval which was adjusted with age, sex.

### Ethical Issues

This study was conducted in full conformance with the ICH E6 guideline for Good Clinical Practice (ICH-GCP) and the principles of the Declaration of Helsinki and ethical clearance from the respective institutional review boards [ERC No-NINS 24-120-21].

## Results

A total of 689 ischemic stroke patients were screened, 156 had confirmed Cardio-embolic stroke. So, the frequency of cardio-embolic stroke in NINS is 22.64%. Overall, the male to female ratio was 1.3:1, mean age of 63 (± 15) years. The most commonly affected age group is 61-70, 45 (28.8%). Sixty-six (42.3%), 17 (12.1%) used to smoke & drinks alcohol accordingly. Hypertension 119 (76.3%), Atrial fibrillation 107 (68.6%) and ischemic heart disease 61 (39.1%) were most common risk factors. However, it is worthwhile to mention 56 (35.9%) had acute myocardial infarction as important risk factors. Interestingly, we found only 23 (14.7%) patients with chronic rheumatic heart diseases, and 11 (7.5%) had hyperthyroidism. Duration of hospital stay was 7 days (minimum 3, maximum 17) in group A while in group B it was 2 days (minimum 1 and maximum 21 days). For Group A, NIH stroke scoring during admission was 9 (minimum 2, maximum 26), while on discharge it was 4 (minimum 0, maximum 11). For Group B, NIH stroke scoring during admission was 21 (minimum 7, maximum 29), while on discharge it was 8 (minimum 4, maximum 11). Most of the cardio-embolic stroke had moderate severity 56 (35.9 %) in Group A while for Group B it was severe 25 (16.1%) [Table 1].

**Table 1.** Demographic Characteristics and severity of Cardio-embolic Patients (n= 156) NIHSS- National Institute of Health Stroke Severity Score COPD- Chronic obstructive lung disease

| Characteristics | Results |  |  |
| --- | --- | --- | --- |
|  | Group A- Alive<br>(109) | Group B- Death<br>(47) | Total (156) |
| **Age ( Mean, SD)** | 61.7 ± 16.6 | 66.7 ± 13.8 | 63.2 ± 15.9 |
| **Age Group ( Years, frequency, percentage)** |  |  |  |
| 21-30 | 2 (1.3) | 1 (0.6) | 3 (1.9) |
| 31-40 | 12 (7.7) | 1 (0.6) | 13 (8.3) |
| 41-50 | 12 (7.7) | 1 (0.6) | 13 (8.3) |
| 51-60 | 26 (16.7) | 13 (8.3) | 39 (25) |
| 61-70 | 29 (18.6) | 16 (10.2) | 45 (28.8) |
| 71-80 | 16 (10.2) | 8 (5.1) | 24 (15.3) |
| 81-90 | 8 (5.1) | 5 (3.2) | 13 (8.3) |
| 91-100 | 4 (2.6) | 2 (1.3) | 6 (3.8) |
| **Sex** |  |  |  |
| Male | 57 (36.5) | 30 (19.2) | 87 (55.8) |
| Female | 52 (33.3) | 17 (10.9) | 69 (44.2) |
| Smoker | 45(28.8) | 21 (13.5) | 66 (42.3) |
| Alcoholic | 12 (7.7) | 5 (3.2) | 17 (10.9) |
| Family H/O Stroke or Heart Disease | 59 (37.8) | 20 (12.8) | 79 (50.6) |
| **Co-morbidities**( frequency, percentage) |  |  |  |
| Hypertension | 79 (50.6) | 40 (25.6) | 119 (76.3) |
| Atrial Fibrillation | 78 (50) | 29 (18.6) | 107 (68.6) |
| Dyslipidemia | 30 (19.2) | 13 (8.3) | 43 (27.5) |
| Acute Myocardial Infarction | 34 (21.8) | 22 (14.1) | 56 (35.9) |
| Diabetes | 39 (25) | 24 (15.4) | 63 (40.4) |
| Ischemic Heart Disease | 40 (25.6) | 21 (13.5) | 61 (39.1) |
| Chronic Rheumatic Heart Disease | 17 (10.9) | 6 (3.8) | 23 (14.7) |
| Chronic Kidney Disease | 9 (5.8) | 9 (5.8) | 18 (11.6) |
| Hyperthyroidism | 11(7.5) | 0 | 11 (7.5) |
| Asthma | 4 (2.6) | 0 | 8 (5.1) |
| COPD | 3 (1.9) | 1(0.7) | 4 (2.6) |
| Prosthetic Heart Valve | 3 (1.9) | 1(0.7) | 4 (2.6) |
| Hospital stay, Median [IQR] | 7 [min 3, max 17] | 2 [ min 1, max 21] | 7 [5-9] |
| NIHSS Score on admission, Median [IQR] | 9 [ min 2, max 26] | 21 [min 7, max 29] | 13 [ 7- 19] |
| NIHSS Score on discharge, Median [IQR] | 4 [ min 0, max 11] | 8 [min 4, max 11] | 5 [3-7] |
| **NIHSS Severity** |  |  |  |
| Mild | 13 (8.3) | 0 | 13 (8.3) |
| Moderate | 56 (35.9) | 4 (2.6) | 60 (38.5) |
| Moderate to severe | 27 (17.3) | 18 (11.5) | 45 (28.8) |
| Severe | 13 (8.3) | 25 (16.1) | 38 (24.4) |
| On Anticoagulant therapy | 106 (67.9) | 37 (23.7) | 143 (91.7) |
| On Beta Blocker | 103 (66) | 33 (21.1) | 136 (87.1) |

Regarding ECG observation, atrial fibrillation was seen in 107 (68.6%) followed by acute myocardial infarction 56 (35.9%). However, in echocardiogram ejection fraction was reduced in 39 (24.8%) although wall motion hypokinesia present in 47 (30.1%) [Table 2].

**Table 2:** Cardiac Abnormality observed in Electrocardiography and Echocardiography (n = 156)

| Findings | Frequency (%) |
| --- | --- |
| Atrial Fibrillation | 107 (68.6) |
| Left Ventricular Hypertrophy | 26 (16.7) |
| STEMI | 33 (21.2) |
| IHD | 31 (12.1) |
| NSTEMI | 23 (19.9) |
| Variable AV Block | 9 (5.7) |
| Atrial Flutter | 5 (3.2) |
| **Echocardiography** |  |
| Rhythm Abnormality | 113 (72.4) |
| Concentric Hypertrophy | 31 (19.8) |
| Valvular Heart disease | 23 (14.7) |
| Valve Replacement | 4 (2.6) |
| Wall Hypokinesia | 47 (30.1) |
| Reduced EF | 39 (24.8) |
| Preserved EF | 116 (75.2) |

STEMI- ST Elevated Myocardial Infarction, NSTEMI- Non ST Elevated Myocardial Infarction, IHD- Ischemic
Heart Disease, AV- Atrio-Ventricular, EF- Ejection Fraction.

Most commonly involved lobe in cardio-embolic stroke was parietal 50 (32.1%) but interestingly parieto-frontal involvement was also common 42 (26.9%). Middle cerebral artery was the most frequently involved vascular territory 116 (74.4%) [Table 3].

**Table 3:**
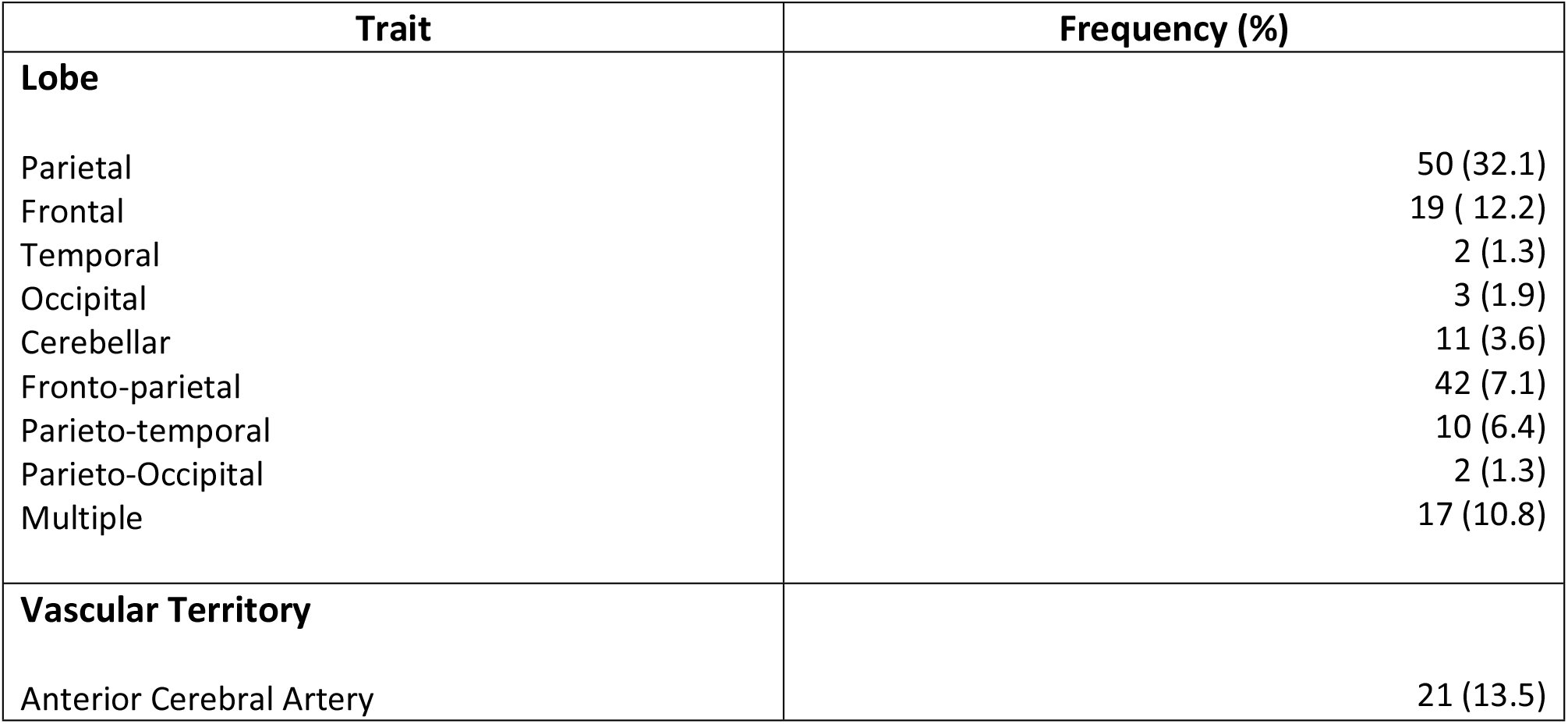
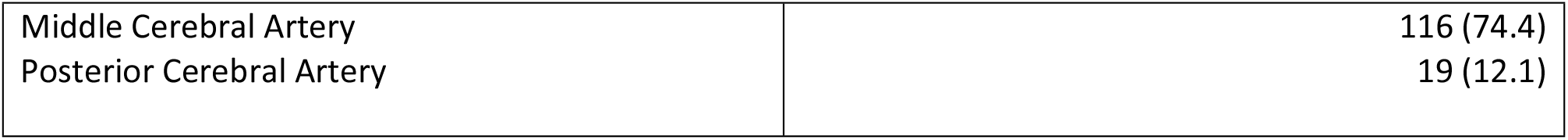
Site of Infarction with involved vascular territory in Brain (n=156)

| Trait | Frequency (%) |
| --- | --- |
| **Lobe** |  |
| Parietal | 50 (32.1) |
| Frontal | 19 ( 12.2) |
| Temporal | 2 (1.3) |
| Occipital | 3 (1.9) |
| Cerebellar | 11 (3.6) |
| Fronto-parietal | 42 (7.1) |
| Parieto-temporal | 10 (6.4) |
| Parieto-Occipital | 2 (1.3) |
| Multiple | 17 (10.8) |
| **Vascular Territory** |  |
| Anterior Cerebral Artery | 21 (13.5) |
| Middle Cerebral Artery | 116 (74.4) |
| Posterior Cerebral Artery | 19 (12.1) |

90 days clinical outcome was variable as modified Rankin score (disability scoring) was 3 (minimum 0, maximum 5), had some symptoms 37 (29.1%) and minor disability 31 (24.4 %) among 109 patients. Hemorrhagic infarct occurred in 29 (18.5%) patients; among them 5 (3.2%) had developed after starting anticoagulant. Only 8 (7.3%) patients developed recurrent ischemic stroke even though on anticoagulant & beta blocker while only 7 (4.5%) suffered hemorrhagic stroke. Overall mortality was 47 (29.9%), among them 30 (19.2%) died on 24-48 hours of hospital admission while 17 (10.9%) within 90 days of hospital discharge [Table 4].

**Table 4:** Outcome of cardio-embolic stroke patients at 90 days of enrollment in study (n=156)

| Outcomes | Group A- alive | Group B-death | Results |
| --- | --- | --- | --- |
| **MRS ( Modified Rankin Score) Median [IQR]**<br>At 3 months | 2 [min 0, max 5]<br>3 [ 2-3] | 6 |  |
| **MRS Severity at 90 days** |  |  |  |
| No limitations | 16 (12.6) | 0 | 16 (12.6) |
| Some Symptoms | 37 (29.1) | 0 | 37 (29.1) |
| Minor disability | 31 (24.4) | 0 | 31 (24.4) |
| Moderate disability | 21 (16.5) | 0 | 21 (16.5) |
| Severe disability | 5 (3.9) | 0 | 5 (3.9) |
| Death |  | 17 (13.4) | 17 (13.4) |
| Hemorrhagic Infarct | 6 (3.8) | 23 (14.7) | 29 (18.5) |
| Anticoagulant induced hemorrhage | 2 (1.3) | 3 (1.9) | 5 (3.2) |
| **Recurrent Stroke within 90 days** |  |  |  |
| Ischemic | 6 (3.8) | 2 (1.3) | 8 (7.3) |
| Hemorrhagic | 3 (1.9) | 4 (2.6) | 7 (4.5) |
| Death |  |  | 47 (29.9) |

### Outcomes Results

Risk factors associated with mortality were acute myocardial infarction OR 1.6 [1.14 – 2.52] p =0.04, severe stroke OR 9.45 [3.57 – 25.03], p <0.001, hypotension OR 3.12 [2.07 – 4.68], **p < 0.001**, raised Troponin-I OR 1.89 [1.16-2.99], **p< 0.01**, raised blood sugar OR 1.82 [1.14 - 2.89], **p < 0.02**, raised Creatinine OR 2.41 [1.52 - 3.84] **p < 0.01, low ejection fraction OR** 3.38 [2.17-5.27] **p < 0.001, large infarct** 53.67 [7.59 - 379.47] **p < 0.001, hemorrhagic infarction OR** 4.43 [2.89 – 6.84] p< 0.001 and aspiration pneumonia OR **1.9 [1.28-2.39]** p = 0.01. [Table5].

**Table 5:** Risk factors associated with mortality among cardio-embolic stroke patients.

| Risk Factors | OR [ 95% CI ] | P value |
| --- | --- | --- |
| Age | 0.98 [0.95-1.01] | 0.08 |
| Sex | 1.40 [0.85-2.32] | 0.25 |
| Smoker | 1.1 [0.68-1.77] | 0.83 |
| Diabetes | 1.50 [0.96-2.48] | 0.11 |
| Hypertension | 1.7 [0.87-3.63] | 0.14 |
| Ischemic heart disease | 1.26 [0.78-2.02] | 0.45 |
| **Chronic kidney disease** | **1.82 [1.06 - 3.10]** | **0.04** |
| Chronic rheumatic heart Disease | 0.85 [0.41 -1.76] | 0.83 |
| Atrial fibrillation | 0.74 [0.46-1.45] | 0.30 |
| **Acute Myocardial infarction** | **1.6 [1.14 – 2.52]** | **0.04** |
| **Raised Troponin** | **1.89 [1.16-2.99]** | **0.01** |
| **Raised Blood Sugar** | **1.82 [1.14 - 2.89]** | **0.02** |
| **Raised Creatinine** | **2.41 [1.52 -3.84]** | **0.001** |
| **Hypotension** | **3.12 [2.07 – 4.68]** | **<0.001** |
| **Severe Stroke** | **9.45 [3.57 – 25.03]** | **<0.001** |
| **Low Ejection fraction** | **3.38 [2.17-5.27]** | **<0.001** |
| **Large Infarct** | **53.67 [7.59 - 379.47]** | **<0.001** |
| **Hemorrhagic Infarction** | **4.43 [2.89 – 6.84]** | **< 0.001** |
| **Aspiration Pneumonia** | **1.9 [1.28-2.39]** | **0.01** |

## Discussion

Cardio-embolic stroke prevalence has been increasing for the last few decades especially among the elderly. Approximately one in four ischemic strokes is of cardio-embolic origin [3], exerting a profound societal impact, association with greater disability, higher mortality rates, and higher treatment costs as compared to patients with strokes from other causes [16,17]. To the best of our knowledge, this is the 1^st^ prospective cohort study that evaluates hospital frequency, clinical outcome, and risk factors determination for mortality among cardio-embolic stroke patients admitted in the largest neurological hospital of Bangladesh. Bhowmik NB et al. described hospital frequency of cardio-embolic stroke as 4.9% but failed to confirm in 17.2% [16], those who were suspected cases for cardio-embolic stroke. Manorenj S et al. [18] observed 11.6% in a prospective study from south India. However, we have observed frequency is 22.05%, it’s obvious there is a significant difference. As we have large sample size, ambulatory 24 hour ECG facility, and portable echocardiogram that helped us for better identification. The prevalence of cardio-embolic stroke in Bangladesh is compatible to the western report of 15-30% [19]. In our cohort study risk factors profile demonstrated hypertension followed by atrial fibrillation are the predominant factors associated with cardio-embolic stroke, very similar to the observation of Manorenj S et al. [18] but different from Henninger N et al [20] who found atrial fibrillation as the predominant factor. Hypertension is certainly the most common condition affecting humans and one in every five Bangladeshi suffers from high blood pressure according to Rahman M et al. [21] from Bangladesh. Cardiac involvement usually occurs in patients with hypertension can lead to the development of atrial fibrillation & stroke [22]. Recent myocardial infarction with left ventricular dysfunction causing aneurysm or thrombus in the heart that can cause cardio-embolic stroke [23]. We do have a similar observation as (28.6%) of our patients suffered acute MI, a majority had low ejection fraction in (23.5%) before the development of stroke, like MacDougall NJJ et al [23]. To identify ischemic stroke was due to cardio-embolic or thromboembolic we went for an MR angiogram of neck vessels and brain in almost every suspicious cases along with an MRI of Brain. Most of the embolic stroke in present cohort involved predominantly left middle cerebral artery with the parietal lobe of the brain which means the source of embolus was the heart. Manorenj S et al. [18], Henninger N et al. [20] & Hart RG [24] had also observed similar vascular & site of brain involvement. Cardiac emboli arising from the cardiac chambers are often large and cause severe stroke, disability, and mortality. They also have high chances of early as well as late embolic recurrences. Hence, early identification of cardio-embolic stroke is crucial for planning the appropriate treatment mode (anticoagulation) and prevention strategies. In our cohort, the majority (45.7%) suffered moderate to severe stroke according to NIHSS grading. Twenty-four (17.1%) patients out of 40 died within 24 hours of hospital admission. Probably large infarct with severe stroke with acute STEMI (ST elevated myocardial infarction) with low ejection fraction (EF < 40%) and delayed referral to specialized stroke unit were the responsible factors behind in-hospital mortality. It is worthwhile to mention that 24 (17.1%) patient’s died during hospital stay where Arboix A & Alió J. mentioned in-hospital mortality in their study was 27.3% [25]. Modified rankin score (MRS≥3) at 90 days was not favorable for those who are alive because most of them have minor to moderate disability. Henninger N et al. [20] in their study also observed an unfavorable 90-day functional status among cardio-embolic patients.

Occluded intracranial vessels with early recanalization and ischemic infarct with hemorrhagic transformations are suggestive of a cardiac-embolic stroke [26]. We observed 21.4% cases of hemorrhagic infarction mostly due to the disease process. However, we also observed anticoagulant-induced hemorrhage in the brain especially those who were on warfarin, had chronic kidney disease, high blood pressure, atrial fibrillation, elderly and labile INR. Hart RG also mentioned those risk factors causing intracerebral hemorrhage in patients who were on warfarin. Nevertheless, even after adequate anticoagulation, few of our patients suffered a recurrent ischemic stroke. Seiffge DJ et al. [28] described during follow-up of 6128 patient’s of acute ischemic stroke with atrial fibrillation on anticoagulant therapy, 289 patients had a further acute ischemic stroke (4.7% per year), 90 patients had ICH (1.5% per year), and 624 patients died (10.2% per year) while on an anticoagulant. Anticoagulation therapy is stills not enough to prevent recurrent ischemic stroke. That’s why further study is needed to find out the pharmacodynamics of different anticoagulants among those who are on an anticoagulant. One of the major concerns of cardio-embolic stroke is long-term mortality. We analyzed those patients who died in our study duration through binary logistic regression model to find out predictors of mortality. We have found hypotension, acute myocardial infarction, low ejection fraction; large infarct, severe stroke, and aspiration pneumonia were significantly associated with mortality, similar to Byun JI et al. [16], Henninger N et al. [20], MacDougall NJJ et al [23]. So, cardio-embolic stroke patients need specialized units run by a multidisciplinary team including intervention facilities and close monitoring to reduce mortality.

Our study has several strengths, like study design was the prospective cohort, having adequate sample size, patients having an ischemic stroke for the first time were included and conducted in a center equipped with adequate investigation facilities, and dedicated team who kept all the patients under regular follow up even in Covid pandemic. However, we do have some limitations. It is a single center study; did not evaluate adequately who suffered a recurrent ischemic stroke within 90 days of hospital discharge although prescribed medications were appropriate. Moreover, we showed some factors that were associated with poor outcome (mortality) through binary logistic regression model but further large-scale study is needed to find out a causal relationship between those factors with mortality.

## Conclusion

To the best of our knowledge, our study is the first-ever work done in Bangladesh to determine the frequency of cardio-embolic stroke in a specialized center, narrated clinical outcomes, determined predictors of mortality, and frequency of anticoagulant induced intracerebral hemorrhage among those stroke patients that most clinician fears off. As mortality in cardio-embolic stroke is high so early identification, timely referral to a specialized hospital, managing those risk factors related to mortality by a multidisciplinary team should be considered as a corner stone in cardio-embolic stroke management. We believe that it is going to modify not only standard protocol in the different centers but also health policy related to cardio-embolic stroke.

## Data Availability

Data set can be shared after acceptance of this article for publications.

## Acknowledgments

We are grateful to our patients and their relatives who gave informed consent for participation in this study. Special thanks to Professor Quazi Deen Mohammad Sir, director of NINS, for his continuous support. Moreover, we are grateful to radiology department for their enormous help regarding radio-imaging of the brain & neck vessels.

## Abbreviations

AF: Atrial fibrillation
COPD: Chronic Obstructive Pulmonary Disease
CRHD: Chronic Rheumatic Heart Disease
ECG: Electrocardiogram
IHD: Ischemic Heart Disease
ICH: Intra Cerebral Hemorrhage
MRS: Modified Rankin Score
MI: Myocardial Infarction
NIHSS: National Institute of Health Stroke Scale
TIA: Transient Ischemic Attack
TSH: Thyroid Stimulating Hormone

